# Karawun: assisting evaluation of advances in multimodal imaging for neurosurgical planning and intraoperative neuronavigation

**DOI:** 10.1101/2021.09.09.21262253

**Authors:** Richard Beare, Bonnie Alexander, Aaron Warren, Michael Kean, Marc Seal, Alison Wray, Wirginia Maixner, Joseph Yuan-Mou Yang

**Affiliations:** Developmental Imaging, Murdoch Children’s Research Institute, Melbourne, Australia; Department of Neurosurgery, The Royal Children’s Hospital, Melbourne, Australia; Neuroscience Research, Murdoch Children’s Research Institute, Melbourne, Australia; Medical Imaging, The Royal Children’s Hospital, Melbourne, Australia; Department of Paediatrics, The University of Melbourne, Melbourne, Australia; Department of Medicine, Monash University, Melbourne, Australia; Department of Medicine, The University of Melbourne, Melbourne, Australia; Epilepsy Neuroinformatics Laboratory, The Florey Institute of Neuroscience and Mental Health, Melbourne, Australia

**Author notes:** Corresponding author: Dr. Joseph Yuan-Mou Yang, The Royal Children’s Hospital and the Murdoch Children’s Research Institute Melbourne, Victoria, Australia.

**Keywords:** Tractography, diffusion imaging, DICOM, neuronavigation

## Abstract

Submitted to *Magnetic Resonance in Medicine*

**Purpose:** To introduce a tool allowing neurosurgeons to evaluate the results of research tractography workflows for presurgical planning and intraoperative image-guidance, using standard neurosurgical navigation platforms.

**Theory and Methods:** Improving communication between neurosurgeons and researchers developing new image acquisition and processing methods is critical for rapid translation of research to surgical practice. Presenting research outputs within existing clinical workflows is one approach that can assist such interdisciplinary communication. Neurosurgical navigation platforms can display and manipulate a wide range of medical image data and associated delineations and thus allow clinicians to evaluate the impact of new imaging research on their work. Currently, it is extremely difficult to integrate research-based image processing outputs into standard neurosurgical navigation platforms.

**Results:** In this note we introduce *Karawun*, an open-source software tool for converting outputs from research imaging pipelines, especially diffusion MRI tractography reconstructions using advanced methodologies currently unavailable on commercial navigation platforms, into forms that can be imported into the *Brainlab* neurosurgical navigation platform (Brainlab AG, Munich, Germany). The externally created tractography images and delineations can be viewed and manipulated as if they were created by *Brainlab*. We illustrate how two surgical workups, created using open-source tools and different processing choices can be presented to the neurosurgeon who can evaluate the impact of the differences between the two workups on surgical decisions.

**Conclusion:** *Karawun* allows researchers developing novel imaging methodologies to display their results in environments that are familiar to clinical end-users, especially neurosurgeons, thus assisting translation of research into clinical practice.

## Introduction

One of the goals of magnetic resonance imaging (MRI) research is improved patient outcomes. The potential benefits to patients offered by advances in the field must be rapidly and clearly communicated to clinical end users if this is to be achieved. However, this is extremely difficult. The MRI research domain is very broad, including fields ranging from physics of hardware and pulse sequence design through to image reconstruction and processing techniques. The application of MRI in clinical practice is similarly broad and complex, involving many clinical specialties, with numerous patient and clinical factors contributing to the choice and interpretation of imaging studies. Communicating the potential implications of MRI research developments is thus extremely challenging. In this note we introduce a tool that can assist communication between MRI researchers and neurosurgeons.

Advanced neuroimage acquisition and analysis techniques are active areas of research that have the potential to enhance neurosurgical planning and intraoperative image-guidance (also known as navigation or neuronavigation), contributing to improved patient care. However, neurosurgeons face a “chicken and egg” problem in that it is difficult to evaluate new imaging methods in a clinical environment while it is impractical for vendors to provide new methods without recommendations from clinicians.

An approach that addresses some of these issues is importing the results of research processing pipelines into standard neuronavigation viewing platforms that are familiar to neurosurgeons, thus allowing evaluation of new results and comparison with existing methods. This has traditionally been a difficult task, but emerging vendor support for new MRI data standards makes it feasible.

In this technical note, we introduce *Karawun*,^1*^ a tool designed to facilitate this approach and thus improve communication between neurosurgeons and imaging researchers, especially for the diffusion MRI tractography outputs that are of particular interest for surgical planning and intraoperative neuronavigation.

## Theory

MRI based imaging techniques are important in presurgical planning and intraoperative guidance for modern neurosurgical practice.^1–3^ Critical information may be derived from routine clinical MRI and advanced neuroimaging research techniques that have demonstrated emerging clinical potential. For example, conventional MRI sequences, such as T1-weighted imaging with and without Gadolinium-contrast, and T2-weighted imaging, are used to reveal where the ideal resection plane lies within the boundary of normal and pathological tissue. Visualization of structural MRI data can be augmented by 3D sulcal-gyral and cortical venous reconstructions, improving the clarity and accuracy of intraoperative patient-specific neuroanatomy, compared to standard 2D MRI display.^4^ When performing surgery within eloquent brain regions, advanced neuroimaging techniques, such as task-based Blood-Oxygenation Level Dependent functional MRI (BOLD-fMRI) and diffusion MRI tractography are utilised to locate eloquent cortex and the associated white matter (WM) tracts.^5–7^ Accurate delineation of the lesion to these eloquent neuronal structures is critical for planning the safe surgical corridors and to ensure functional preservation.

Methods of acquiring and processing diffusion MRI and tractography reconstructions have substantially advanced over the past two decades.^3,8–12^ Every step involved in generating the tract images has benefited. Advances in MRI hardware and sequences has enabled acquisition of DWI appropriate for advanced modelling within clinically feasible times.^13^ Models that estimate multiple WM fiber orientations per MRI voxel^14–17^ and related advances in tractography algorithms^18–20^ are now well recognised to overcome many limitations of the conventional diffusion tensor imaging (DTI) model^21,22^ and a deterministic tracking algorithm, such as the Fiber Assignment by Continuous Tracking (FACT) algorithm.^23^ This facilitates more robust tractography reconstructions better representing underlying WM tract anatomy. Many of these methods are implemented in open source and/or freely available software packages supported by active communities of developers and users.^24^

Thus, the wide range of neuroimaging software tools created by the research community represents an enormously powerful and flexible resource that can be used to develop and test new acquisition and processing approaches. However, clinical uptake of these advanced neuroimaging tools is very limited.^3,25^ It remains very difficult for clinicians to evaluate the improvements to clinical care that they may offer. In part this is because the research software outputs do not match the file format required by commercial surgical neuronavigation software platforms used in clinical practice.

In this note we demonstrate a comparison procedure using *Karawun* to import two neurosurgical workups into *Brainlab* (Brainlab AG, Munich, Germany). *Karawun* transforms results of an imaging workup generated using open-source tools, including 3D segmentation data of WM tracts and tumor mask, and converts them to the specialised Digital Imaging and Communications in Medicine (DICOM) format supported by commercial vendors. Recent extensions to the DICOM standard include support for a range of quantitative forms of image data, including segmentation objects, parametric images that are intended to allow interoperability between platforms, and *Karawun* uses these standards. The converted data can then be imported into a familiar visualization environment, thus providing neurosurgeons with the ability to evaluate imaging workups and investigate clinical impact of changes to components of imaging workflows.

These multimodal imaging-based workflows can be established using a wide range of software tools created by the research community. There are many choices, and it is not our aim to describe the specifics. Instead, we will summarize the common steps and typical outputs.

### Typical components of imaging workup

A sample workflow is illustrated in Figure 1

**Figure 1:**
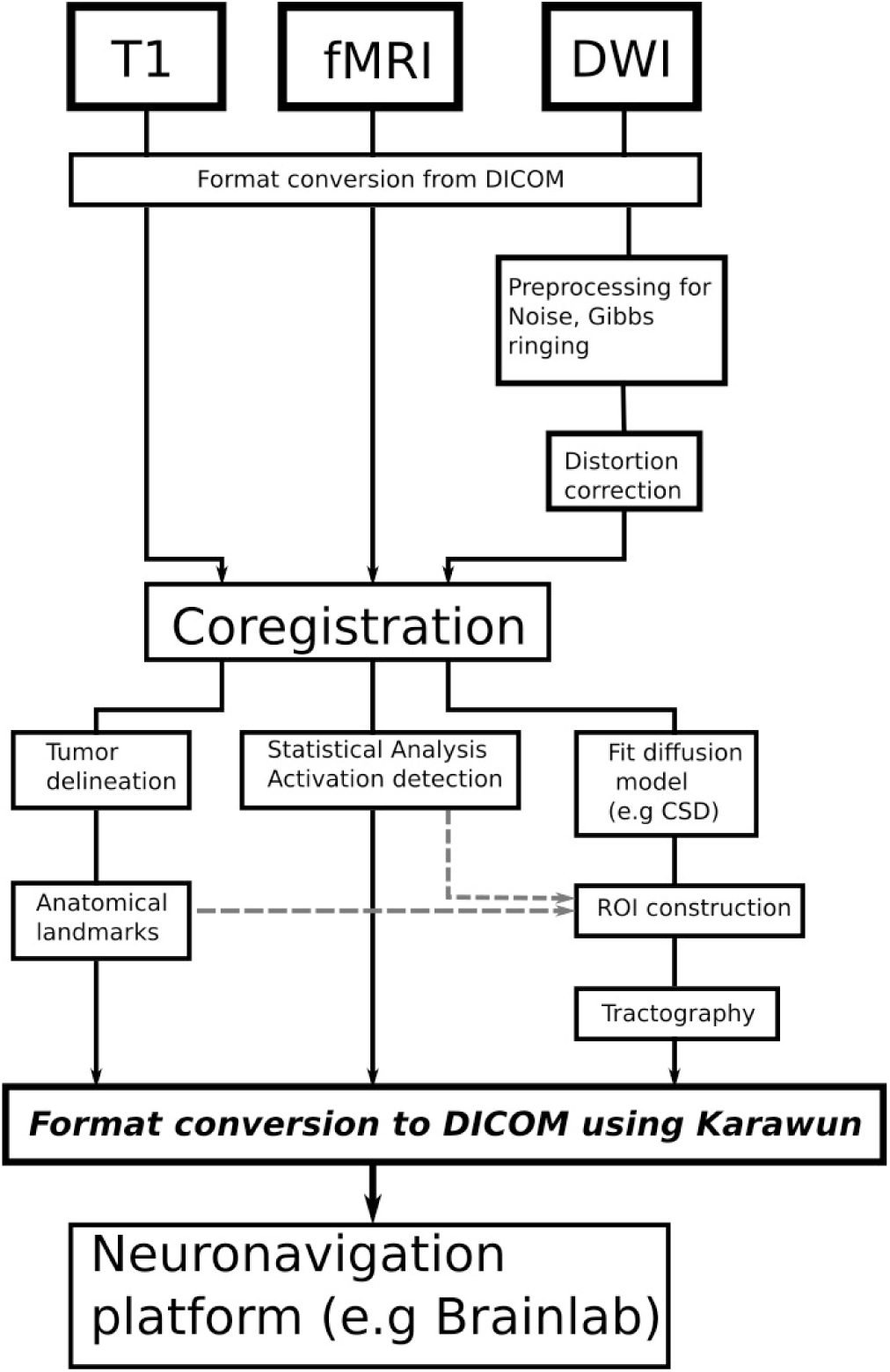
A typical workflow for neurosurgical planning using structural, functional and diffusion MRI data.

#### Format conversion

The typical first step in many research software tools is converting the raw MRI data into more analyzable file formats (e.g the Nifti format).

#### Coregistration

Scans of the same patient acquired at different times, and across different imaging modalities are aligned with each other, or “coregistered”. This process allows features visible in one scan type to be projected onto another. For example, regions of activation derived from BOLD-fMRI may be aligned with a T1-weighted scan.

#### Statistical analysis

Many forms of imaging data require statistical analysis. Task-based BOLD-fMRI data typically undergo statistical analyses to determine areas of task-related BOLD signal changes. The output is a 3D statistical map conveying the association between areas of brain activation and the task.

#### Delineation of regions of interest

Regions of interest (ROIs) or masks are used to visualize critical components or provide information to other processing steps. Surgical targets may be delineated manually with a drawing tool. Automated procedures, such as implemented by FreeSurfer (https://surfer.nmr.mgh.harvard.edu/), can parcellate a brain into anatomical regions. Vascular structures may be delineated by applying a threshold to a Gadolinium-contrast enhanced image.

#### Tractography

A tractography image contains a set of *streamlines*. Streamlines are generated from DWI data using a complex processing pipeline, including data noise reduction and image distortion correction before fitting a model to estimate WM fiber orientation. Model estimation is followed by tractography reconstruction based on a tracking algorithm, which may be either a deterministic or probabilistic method. Tractography reconstructions are guided by using ROIs defined based on known WM tract anatomy. Each of these steps may be performed by several different tools in different ways that may lead to different results, and it is important for neurosurgeons to evaluate the impact of such differences on clinical decisions.

### Typical outputs of imaging workup

The classes of data produced by an imaging workup relevant to this note are:

- A set of coregistered anatomical images - for example, T1- and T2-weighted structural images.
- A set of tract files, each containing one or more 3D models of WM tracts generated by a diffusion MRI tractography pipeline.
- A set of mask images, also known as segmentation/labelled images. Masks can be converted to 3D models by visualization software. Masks are used to represent the tumor and BOLD-fMRI activation regions in this note.

*Karawun* is designed to convert such datasets to DICOM. Anatomical images are converted to a traditional DICOM image format and use a common reference frame. Tracts and masks are converted to DICOM quantitative imaging formats based on the DICOM Segmentation Information Object Definition. Quantitative imaging formats are designed to support the exchange and visualization of data derived from image data, such as segmentations, and are becoming more widely implemented and supported.^26–29^

## Methods

### Software structure

*Karawun* is implemented as a python 3.7 package and instructions for installation and use are available at https://github.com/DevelopmentalImagingMCRI/karawun. It can process 3D Nifti image files and tract files produced by the MRtrix3 software suite (www.mrtrix.org).^24^ Participant-specific metadata is copied from the original DICOM (e.g., participant name, date of imaging acquisition, etc). Workup specific metadata, such as file names, are inserted into DICOM fields and are visible in the Brainlab interface.

### Ethics Statement

The project received ethical approval from The Royal Children’s Hospital Melbourne Human Research Ethics Committee (HREC) and received governance authorisation at the Melbourne Children’s Campus (incorporating The Royal Children’s Hospital, Murdoch Children’s Research Institute and the University of Melbourne Department of Paediatrics). HREC approval date: 18 March 2021. HREC Reference Number: HREC/72907/RCHM-2021

The project used retrospectively collected clinical data and aim 1 included reporting the surgical outcomes of neurosurgical cases that had used data imported by *Karawun* for planning and intraoperative guidance.

### Participant

A child aged 10-15 presented with a large left parieto-occipital brain tumor revealed on diagnostic MRI (Figure 2). No language, motor or visual deficits were detected on formal neurological and functional examinations. An open craniotomy and surgical debulk of this tumor was performed, followed by cranial irradiation therapy and chemotherapy. The right hand motor functional cortex, left corticospinal tract and the left optic radiation were mapped to establish a safe surgical corridor and trajectory.

**Figure 2:**
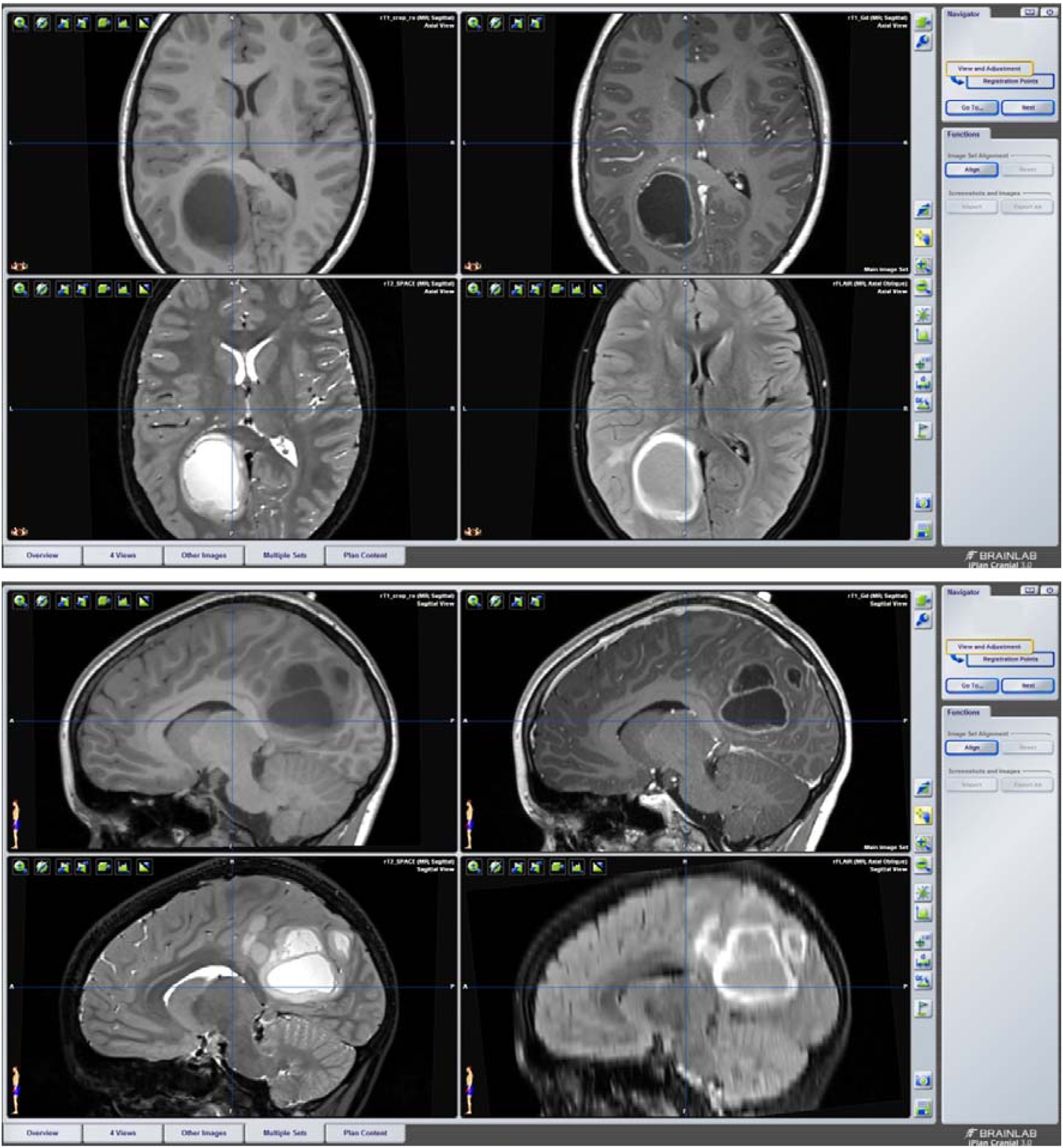
BrainLab iPlan Cranial views showing the left parieto-occipital tumor using conventional clinical MRI sequences – T1-weighted, T1-weighted with Gadolinium contrast, T2-weighted and FLAIR.

### MR Imaging

Presurgical MRI was performed on a 3Tesla Siemens MAGNETOM Prisma scanner with a 64-channel head coil receiver. The following sequences were acquired: volumetric T1-weighted with and without Gadolinium contrast, T2-weighted and non-volumetric FLAIR, multi-shell DWI (b=3000/2000/1000s/mm^2^, 60/45/25 directions) and a right index finger tapping task-based motor BOLD-fMRI scan (standard block design paradigm: 15 secs active finger-tapping blocks, alternating with 15 secs rest blocks, totalling 195 secs).

#### Image processing

##### Coregistration

The T1-weighted images were linearly coregistered to the distortion corrected b0 volume of the DWI data using FMRIB Linear Registration Tool (FLIRT) from FSL (version 6, FMRIB’s Software Library; www.fmrib.ox.ac.uk/fsl).^30,31^

##### Probabilistic multi-tissue CSD and deterministic DTI tractography processing

All DWI processing and tractography reconstruction were performed using MRtrix3.^24^

A multi-tissue CSD technique was used to estimate tissue response function and to model WM fiber orientations.^32^ Tractography was performed using an iFOD2 probabilistic tracking algorithm.^24^ Corticospinal tract and optic radiation tractography were reconstructed based on our previously published methods.^33,34^

An additional pair of the same corticospinal tract and optic radiation was generated for comparison purposes using the same sets of tracking ROIs, and using a DTI model estimated from the b=1000 mm/s^2^ shell of the multishell DWI data, and tractography using the FACT algorithm implemented in MRtrix3.^21,22^ The DTI/FACT combination is the same tractography tools implemented in Brainlab.

##### FMRI processing

A standard block design analysis of the motor finger-tapping task was performed using FSL-FEAT (FMRI Expert Analysis Tool) Version 6.00.^35^

##### Tumor delineation

A tumor mask was delineated manually using the *mrview* visualization tool.

### Brainlab-readable file format conversion using *Karawun*

The set of aligned nifti anatomical files, tract files in MRrtrix3 “.tck” format, and Nifti label images were converted to DICOM image and quantitative formats with the following command:

~~~
importTractography --dicom-template path/to/a/dicom --nifti
T1.nii.gz T1Gd.nii.gz T2.nii.gz Flair.nii.gz --tract-files
left_cst_csd.tck left_cst_dti.tck left_or_csd.tck left_or_dti.tck --
label-files tumor_mask.nii.gz movement_activation_mask.nii.gz -o
path/to/output/folder
~~~

`dicom_template` : an original patient DICOM providing patient details.

`nifti` : a set of anatomical images to be converted to standard DICOM format.

`label-files` : a set of mask images that are converted to DICOM voxel segmentation images.

`tract-files` : a set of MRtrix3 format tractography files that are converted to the DICOM fiber segmentation format.

`-o` : output folder.

### Clinical visualization

Brainlab’s web interface was used to import the *Karawun* output. Combined visualizations of anatomical and processed BOLD-fMRI and diffusion MRI tractography data were created and displayed in Brainlab iPlan cranial. Tractography reconstructed using the two different modeling and tractography techniques were visually compared.

## Results

The Brainlab iPlan cranial neuronavigation platform view of the clinical case example is shown in Figure 3. The two figure panels demonstrate streamline objects representing the left corticospinal tract and the left optic radiation, colored by streamline orientation, and segmentation objects displaying the tumor (purple) and areas of BOLD-fMRI motor activation (red). These objects are overlaid on the coregistered T1-weighted image in this example, although may be overlaid on any other scans that were imported. The tumor and BOLD-fMRI activation are visible as solid objects in the 3D view, and as outlines in the slice views.

**Figure 3:**
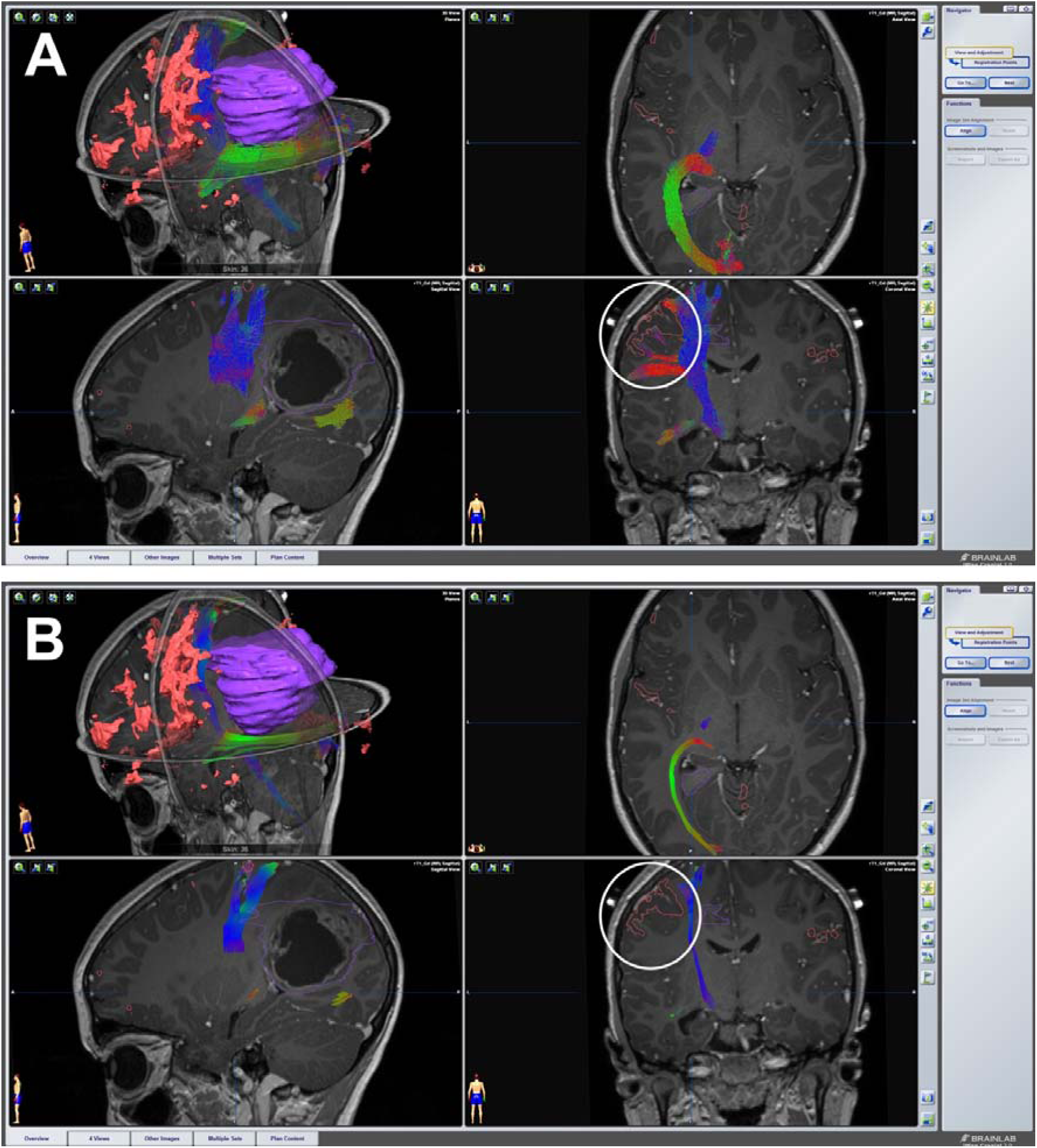
BrainLab iPlan Cranial views of two imaging workups of the same patient using different tractography methods. Manually delineated tumor in purple and motor functional MRI activation in red. Panel A: multi-shell, multi-tissue-Constrained Spherical Decon

The neuronavigation software allows clinicians to explore important spatial relationships between the tumor, the adjacent WM tracts, areas of peri-tumoral motor BOLD-fMRI activation, and anatomical features visible in the background MRI images.

Panel A shows tractography reconstructed using a method combining the multi-tissue CSD model and a probabilistic tracking algorithm, available in MRtrix3. Panel B shows the same tracts reconstructed using the conventional DTI modeling and FACT tracking algorithm. Larger gaps between the tumor margins and both WM tracts are visible in Panel B than A. In Panel B, there is a failure to reconstruct the lateral face-motor projections of the corticospinal tract and lack of spatial overlap between activated finger-motor cortex and the tract, as indicated by the white circles. Inadvertent injuries to these WM tracts due to such false-negative tracking problems will likely result in postoperative permanent motor and/or visual field deficits. For this patient, the resection margins were informed by multi-tissue CSD-based tract objects (i.e. images from Panel A) and there were no transient or permanent functional deficits following surgery.

## Discussion

The translation of advanced neuroimaging techniques into clinical practice is challenging because clinicians, including neurosurgeons, don’t have the opportunity to apply and evaluate the results of these new methods in their normal day-to-day practice.

Our proposed open-source conversion tool, *Karawun*, is designed to bridge this evidence-practice gap by enabling neurosurgeons to have direct means to evaluate the advanced neuroimaging outputs in a familiar clinical software environment. It also serves as a methodological platform for researchers to evaluate research developments that are useful in a clinical environment.

Several brain tumor cohort studies demonstrated improved anatomical plausibility of the tractography reconstruction using advanced multi-fiber models and probabilistic tracking algorithms compared with the deterministic DTI technique from existing surgical neuronavigation devices.^36–38^ The benefits of these more advanced tractography techniques were most pronounced when reconstructing tracts near to the pathology, and in presence of perilesional WM edema.^39^ Importantly, the majority of these studies were conducted with a research focus without implementing clinical translation pathways. With the introduction of *Karawun*, the clinical translation to a selected neuronavigation device is possible.

*Karawun* continues the tradition of open-source software supporting MRI research. We believe that it adds a clinically valuable new option to the suite of open-source software supporting the new DICOM standards.

## Conclusions

This note has introduced a tool that we hope will reduce barriers between researchers developing MRI acquisition and processing techniques and neurosurgeons, who are the eventual consumers of these research efforts. Research groups may use the tool to directly involve neurosurgeons and other clinicians in the evaluation of new workflows and assessment of changes to any part of an existing workflow. Clinicians may more easily explore the impact of techniques that are not (widely) available in clinical suites and investigate the use of advanced acquisitions that are not supported by commercial clinical tools.

## Data Availability

Karawun runs on Windows, OSX and Linux and is freely available via GitHub (https://github.com/DevelopmentalImagingMCRI/karawun). Sample data appropriate for testing purposes is included.

## Data availability statement

*Karawun* runs on Windows, OSX and Linux and is freely available via GitHub (https://github.com/DevelopmentalImagingMCRI/karawun). Sample data appropriate for testing purposes is included.

## Author contributions

Conception and design: Beare, Maixner, Yang. Acquisition of data: Kean, Wray. Analysis and interpretation of data: Alexander, Beare, Warren, Yang. Drafting the article: Beare, Yang. Critically revising the article: all authors. Reviewed submitted version of manuscript: all authors. Approved the final version of the manuscript on behalf of all authors: Yang.

## Acknowledgements

This research was conducted within the Department of Neurosurgery, The Royal Children’s Hospital, and the Developmental Imaging Group, Murdoch Children’s Research Institute at the Melbourne Children’s MRI Centre, Melbourne, Victoria. It was supported by The Royal Children’s Hospital Foundation, Murdoch Children’s Research Institute, The University of Melbourne, Department of Paediatrics, and the Victorian Government’s Operational Infrastructure Support Program.

## Footnotes

1* Karawun is a grass used as a fiber crop by Aboriginal people in southeastern Australia.

